# Acute and longer-term psychological distress associated with testing positive for COVID-19: longitudinal evidence from a population-based study of US adults

**DOI:** 10.1101/2021.03.25.21254326

**Authors:** Michael Daly, Eric Robinson

**Author notes:** Corresponding author: Michael Daly Address correspondence to: Michael Daly Ph.D., Department of Psychology, 1.1.7 Education House, Maynooth University, Maynooth, Ireland.

## Abstract

**Background:** The novel coronavirus (SARS-CoV-2) has produced a considerable public health burden but the impact that contracting the disease has on mental health is unclear. In this observational population-based cohort study, we examined longitudinal changes in psychological distress associated with testing positive for COVID-19.

**Methods:** Participants (N = 8,002; Observations = 139,035) were drawn from 23 waves of the Understanding America Study, a nationally representative survey of American adults followed-up every two weeks from April 1 2020 to February 15 2021. Psychological distress was assessed using the standardized total score on the Patient Health Questionnaire-4 (PHQ-4).

**Results:** Over the course of the study 576 participants reported testing positive for COVID-19. Using regression analysis including individual and time fixed effects we found that psychological distress increased by 0.29 standard deviations (*p* <.001) during the two-week period when participants first tested positive for COVID-19. Distress levels remained significantly elevated (*d* = 0.16, p <.01) for a further two weeks, before returning to baseline levels. Coronavirus symptom severity explained changes in distress attributable to COVID-19, whereby distress was more pronounced among those whose symptoms were more severe and were slower to subside.

**Conclusions:** This study indicates that testing positive for COVID-19 is associated with an initial increase in psychological distress that diminishes quickly as symptoms subside. While COVID-19 may not produce lasting psychological distress among the majority of the general population it remains possible that a minority may suffer longer-term mental health consequences.

## Introduction

The COVID-19 pandemic radically changed daily life for much of the world’s population and by March 2021 had led to 122 million confirmed infections worldwide including almost 30 million in the United States (WHO, 2021). While there has been a surge in research examining the potential population mental health consequences of living through the pandemic (Robinson, Sutin, Daly, & Jones, 2021; Salari et al., 2020), few studies have specifically examined the mental health impact of contracting the novel coronavirus (SARS-CoV-2). A review of studies examining severe acute respiratory syndrome (SARS) and Middle East respiratory syndrome (MERS) concluded that severe coronavirus infections are linked to elevated depression and anxiety disorder in the months following infection (Rogers et al., 2020). In the context of COVID-19, there is growing concern that the disease may lead to prolonged effects after recovery including headaches, muscle and body ache, and persistent tiredness (Sudre et al., 2021) and that these post-acute COVID-19 symptoms (often termed “long COVID”) may result in prolonged mental health problems (Del Rio, Collins, & Malani, 2020). However, there is currently a dearth of evidence on the mental health impacts of contracting COVID-19 and the present study aimed to address this gap by examining longitudinal data on US adults to estimate changes in psychological distress in response to contracting COVID-19.

Research to date suggests that reporting a positive test or symptoms consistent with COVID-19 is associated with raised anxiety and depression levels in cross-sectional samples of the general population in the UK and Ireland (Hyland et al., 2020; Shevlin et al., 2020). Similarly, reporting symptoms compatible with COVID-19 has been linked to worse mental health in a large-scale study (N = 69,054) of university students in France (Wathelet et al., 2020). A large-scale cross-sectional study of UK adults (N = 44,775) has shown that reporting a diagnosis of COVID-19 is associated with raised levels of self-harm and suicidal ideation (Iob, Steptoe, & Fancourt, 2020). Although these studies link potential COVID symptoms or diagnosis with increased mental health problems, the cross-sectional nature of study means that it is plausible that findings are explained by confounding bias or reverse causality. For example, individuals with existing mental health problems may be more likely to report experiencing COVID-19 symptoms or to contract COVID-19 (van der Meer et al., 2020).

In line with this, a large-scale record linkage study of 61 million US adults found that those with a recent diagnosis of a mental disorder were at increased risk of COVID-19 infection (Wang, Xu, & Volkow, 2021). This finding was confirmed by a subsequent study of the health records of almost 70 million patients in the US (Taquet, Luciano, Geddes, & Harrison, 2021). Within the same study, 62,354 patients contracted COVID-19 between January and August 2020 and this group were at an increased risk of first-time diagnosis of psychiatric disorder (particularly anxiety disorder and insomnia), within 3-months relative to those experiencing other health events such as influenza (Taquet et al., 2021). This finding suggests that contracting COVID-19 may increase risk of mental health problems.

However, in the absence of longitudinal data on mental health symptoms prior to infection and data on the background characteristics of patients it is unclear whether this association could be attributed to pre-illness factors, such as subclinical mental health symptoms or confounding lifestyle factors that predispose individuals towards more severe COVID-19 outcomes and poorer mental health (Popkin et al., 2020; Wang, Kaelber, Xu, & Volkow, 2021). It is also not clear whether the increased risk of psychiatric problems identified may be representative of the impact of COVID-19 in the general population. For instance, a follow-up study drawing on the same health records showed that the onset of mood and anxiety disorders at six months was increased only among a subset of COVID-19 patients experiencing brain dysfunction as a result of severe infection (Taquet, Geddes, & Harrison, 2021).

To understand the potential impact of COVID-19 infection on mental health in the general population there is a need for representative longitudinal data which assesses whether pre-COVID-19 mental health symptoms change sequentially as a result of infection. The objective of the current study was therefore to examine longitudinal changes in psychological distress both during infection and after recovery in large nationally representative cohort of US adults. We also examined if results were consistent across population demographics and the extent to which changes in psychological distress were explained by the severity and duration of COVID-19 symptoms (e.g. respiratory difficulties) experienced as a result of infection.

## Methods

### Sample

Participants were drawn from the Understanding America Study (UAS), a nationally representative probability-based longitudinal study of 9063 individuals (Alattar Messel, & Rogofsky, 2018; Kapteyn et al., 2020). The sample was recruited via address-based sampling from the US Postal Service Computerized Delivery Sequence file (Alattar et al., 2018). The UAS surveys are administered via the internet and eligible participants without computers or internet access are provided with internet-connected tablets. From April 1 2020, respondents were invited to take part in a continuous tracking study where participants completed surveys every two weeks during the COVID-19 pandemic. The survey has been used to examine changes in mental health (Daly & Robinson, 2020) and COVID-19 related perceptions and protective behaviors (Crane, Shermock, Omer, & Romley, 2021; Robinson & Daly, 2020).

From a total number of 8,129 participants and 142,573 observations across 23 waves of data collection we excluded 127 participants and 3,534 observations with missing data on the PHQ-4 (Obs. = 2,131) and reported demographic characteristics and COVID-19 test results and symptoms (Obs. = 1,407). We utilize data from the remaining 8,002 UAS panel members who provided a total of 139,035 observations from April 1 2020 to February 15 2021. The average number of participants in each survey wave was 6,045 and participants completed 17.4 out of 23 possible surveys on average. The dates of data collection and number of participants in each survey wave can be seen in Table S1.

Further details of the UAS survey administration and sampling methodology can found elsewhere (Alattar et al., 2018; Kapteyn et al., 2020). The UAS was approved by the University of Southern California human subjects committee internal review board (IRB) and informed consent was obtained from all participants.

### Measures

### Psychological distress

Psychological distress was gauged using the validated four-item Patient Health Questionnaire (PHQ-4), which has been shown to be reliable and responsive to changes in mental health (Kroenke, Baye & Lourens, 2019; Kroenke, Spitzer, Williams, & Löwe, 2009; Löwe et al., 2010). The scale is comprised of two items from the PHQ-9 and two items from the Generalized Anxiety Disorder-7 (GAD-7). The items selected assess core symptoms of anxiety (i.e. “feeling nervous, anxious, or on edge” and “not being able to stop or control worrying”) and depression (i.e. “little interest of pleasure in doing things” and “feeling down, depressed, or hopeless”). Participants were asked how often over the last 2 weeks they have been bothered by these problems and reported their answers on a four-point scale with response options *not at all*□=□0, *several days*□=□1, *more than half of days*□=□2, and *nearly every day* □=□3. Total scores on the scale range from 0 to 12 with higher scores indicating greater distress (mean Cronbach’s *α* = 0.92, range = 0.88 - 0.93). Total PHQ-4 scores were standardized to have a mean of zero and standard deviation of one.

### COVID-19 testing

As part of each survey wave participants reported whether they had been tested for coronavirus since they last completed the UAS continuous tracking survey and were reminded of the date of their most recent survey. Participants reported the result of the coronavirus test using four options: (1) “I have been tested and I tested positive (I had coronavirus)”, (2) “I have been tested and I tested negative (I did not have coronavirus)”, (3) “I have been tested and I do not know the result”, and (4) “I have not been tested”. Those who tested positive were classified as having COVID-19 and others were classified as not testing positive. Where participants reported testing positive in more than one survey wave (typically in waves directly following the first positive COVID-19 test) we examined the first positive test only.

### COVID-19 symptoms

In each survey wave participants indicated whether or not they had experienced common symptoms of COVID-19 in the past 7 days. In line with a recent large-scale assessment of the clinical spectrum of COVID-19 symptoms (Eythorsson et al., 2020) we included assessments of upper respiratory (i.e. runny or stuffy nose, sore throat, sneezing, lost sense of smell), lower respiratory (i.e. chest congestion, cough, shortness of breath), gastrointestinal (i.e. vomiting, diarrhea, abdominal discomfort), and generalized symptoms (i.e. fever or chills or body temperature higher than 100.4 F or 38.0 C, headache, muscle or body aches). The percentage of all symptoms reported in the past 7 days was examined as our primary indicator of participant experiences of COVID-19 (ranging from 0 = no symptoms experienced, to 100 = all symptoms experienced). The composite score of the percentage of symptoms reported demonstrated adequate reliability across survey waves (mean Cronbach’s *α* = 0.76, range 0.72 – 0.79).

### Demographic characteristics and other potential moderating factors

We examined a set of demographic factors that may moderate the association between COVID-19 and psychological distress: participant age (18-39, 40-59, 60+ years), gender (male, female), race/ethnicity (White, Hispanic, Black, Other race/ethnicity), and annual household income levels (≤$40,00, $40,000–$100,000 ≥$100,000 per annum). In addition, we tested whether the potential association between COVID-19 and distress levels was moderated by the presence of a pre-existing diagnosis of a chronic physical health condition (i.e. diabetes, cancer, heart disease, kidney disease, asthma, chronic lung disease, autoimmune disease) or a mental health condition (i.e. anxiety disorder, attention deficit hyperactivity disorder, bipolar disorder, eating disorders, depressive disorders, obsessive compulsive disorder, posttraumatic stress disorder, or schizophrenia/psychotic disorder).

### Statistical analysis

All analyses were carried out in Stata version 15 using survey weights to generate nationally representative estimates. In the UAS sampling weights adjust for differential probabilities of selection into the UAS and post-stratification is used to adjust the weights so that each survey matches the distribution of demographic characteristics of the US population. Further details of the UAS weighting methodology can be found in Angrisani Kapteyn, Meijer, and Saw (2019).

In preliminary analyses, to test the validity of the self-reported COVID-19 positive test data we compared the estimated population prevalence of positive tests in each wave of the UAS and in the US adult population over the same two-week periods from April 1 2020 to February 15 2021. Total case numbers per day were drawn from the Centers for Disease Control and Prevention COVID Data Tracker (CDC, 2021) and an adjustment was applied to correct for the presence of those under 18 in the daily case figures (see Supplementary Materials Section 1). We also compared the prevalence of each COVID-19 symptom examined in this study with the symptoms identified in two large-scale studies examining the symptoms of those with confirmed positive COVID-19 tests (Eythorsson et al., 2020; Wohl et al., 2021). Finally, we tested whether testing positive for COVID-19 was linked to subsequent drop-out or a reduced level of participation in the UAS continuous tracking survey.

Our main analysis used fixed effects regression to estimate the link between reported coronavirus disease and changes in psychological distress within the same individuals across multiple time-points. The basic fixed effects model estimating the association between testing positive for COVID-19 (β_*1*_*COVID*_*it*_) and psychological distress (*PHQ*_*it*_) experienced by individuals *i* across survey waves *t* can be expressed as:

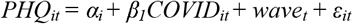

The inclusion of individual fixed effects (*α*_*i*_) provides a control for all stable individual characteristics that could influence susceptibility to COVID-19 or psychological distress (e.g. gender, race, family background). Although individual fixed effects adjust for all unchanging characteristics that could impact this relationship, it remains possible that the link between COVID-19 and distress could be impacted by seasonal and other calendar effects associated with the pandemic (e.g. changing macroeconomic conditions). For this reason, we also included survey wave fixed effects (*wave*_*t*_) for each two-week period during which the study surveys were completed. By including individual and time fixed effects, we eliminated bias from all factors that are fixed over time for individuals and factors that vary over time but are constant across individuals.

The fixed effects approach employed in this study compares the distress levels of an individual when they have tested positive for COVID-19 (and in waves before/after testing positive where examined) with their distress level in other waves and pools this information across the sample. In this way the increase in distress associated with testing positive for COVID-19 can be estimated relative to a “baseline” level (Clark, Diener, Georgellis, & Lucas, 2008), which is the predicted level of distress at other time-points experienced by those who at some stage test positive for COVID-19. For modelling purposes, the sample included both those who were and were not diagnosed with COVID-19 over the course of the study. We also adopted this approach to examine changes in mental health experienced by those who went for a COVID-19 test and tested negative for the disease.

Lags and leads in the effect of COVID-19 were examined to identify whether distress increased before or after testing positive for the disease. Lead effects identified whether distress increased from baseline levels in advance of a positive test. Lag effects identified whether there was a residual effect of COVID-19 on psychological distress in subsequent survey waves after the participant tested positive for the disease. Lag and lead analyses relied on survey waves completed immediately before and after the wave where participants tested positive for COVID-19. In addition, we tested whether the association between COVID-19 and psychological distress was moderated by participant sociodemographic characteristics and the presence of chronic physical or mental health conditions.

Finally, we utilized the fixed effects model outlined above to examine the association between testing positive for COVID-19 and experiencing symptoms of the disease, and the association between COVID-19 symptoms and psychological distress. Drawing on this information we conducted mediation analysis using the *khb* command in Stata to estimate the indirect effect of COVID-19 on psychological distress via COVID-19 symptoms (Karlson, Holm, & Breen, 2012).

## Results

### Sample characteristics

The demographic characteristics and symptoms experienced by those who tested positive for COVID-19 and the remaining UAS participants are shown in Table 1. Those who tested positive were more likely to be aged 40-59 years compared to the remainder of the sample. Those reporting a positive test were also more likely to be of Hispanic race/ethnicity. We found that those who tested positive for COVID-19 did not complete a significantly different number of surveys (M = 17.85 waves completed, SD = 6.5) to participants who did not test positive for COVID-19 (M = 17.34 waves completed, SD = 7.24) (B = 0.51, SE = 0.31, p = .10). Those who tested positive for COVID-19 at any stage in 2020 (N = 426) were significantly more likely than other participants to take part in the UAS survey in 2021 (OR = 1.82, [95% CI: 1.28-2.61], p < .01), though retention was very high for both groups (92% vs. 86.3%).

**Table 1.**
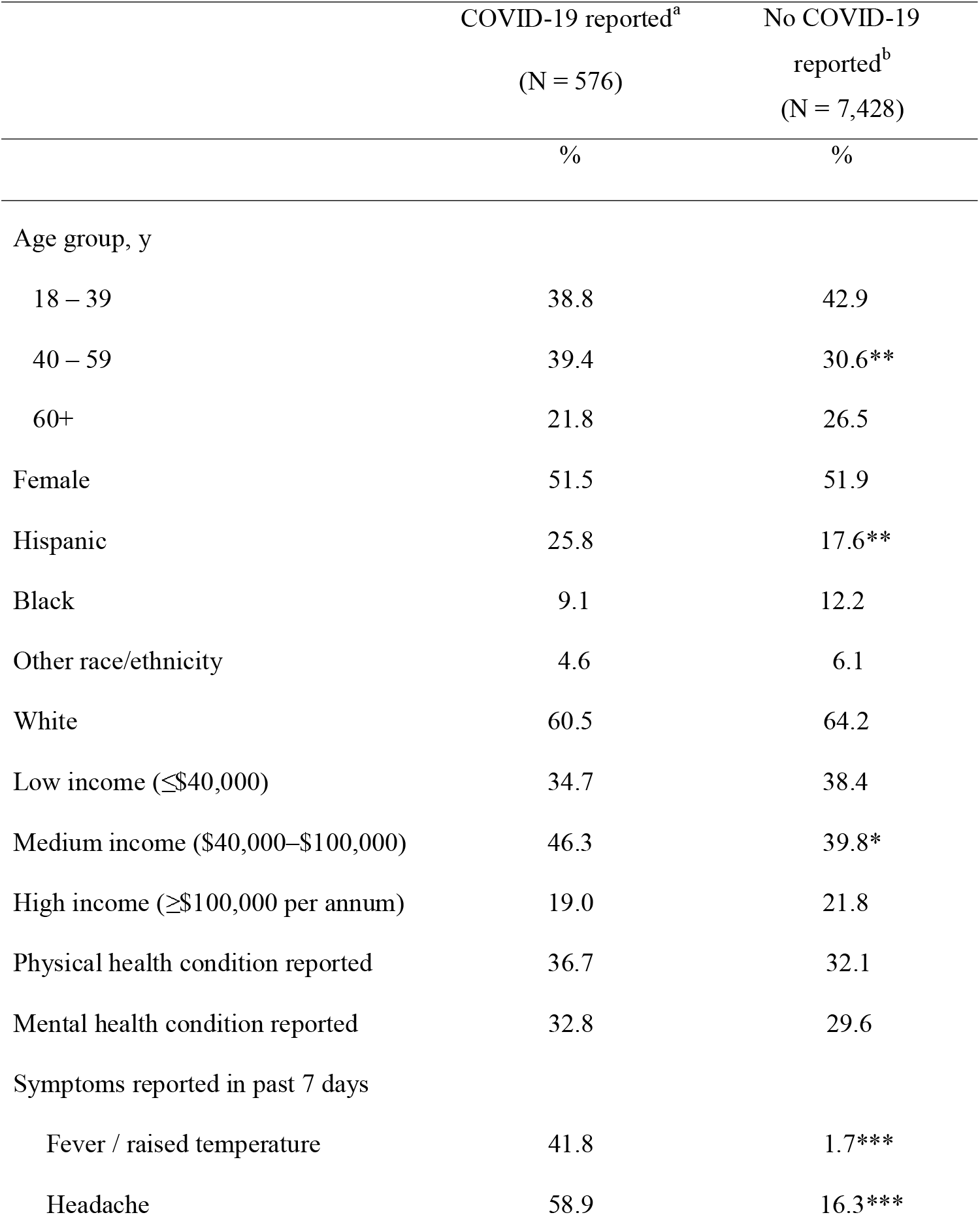
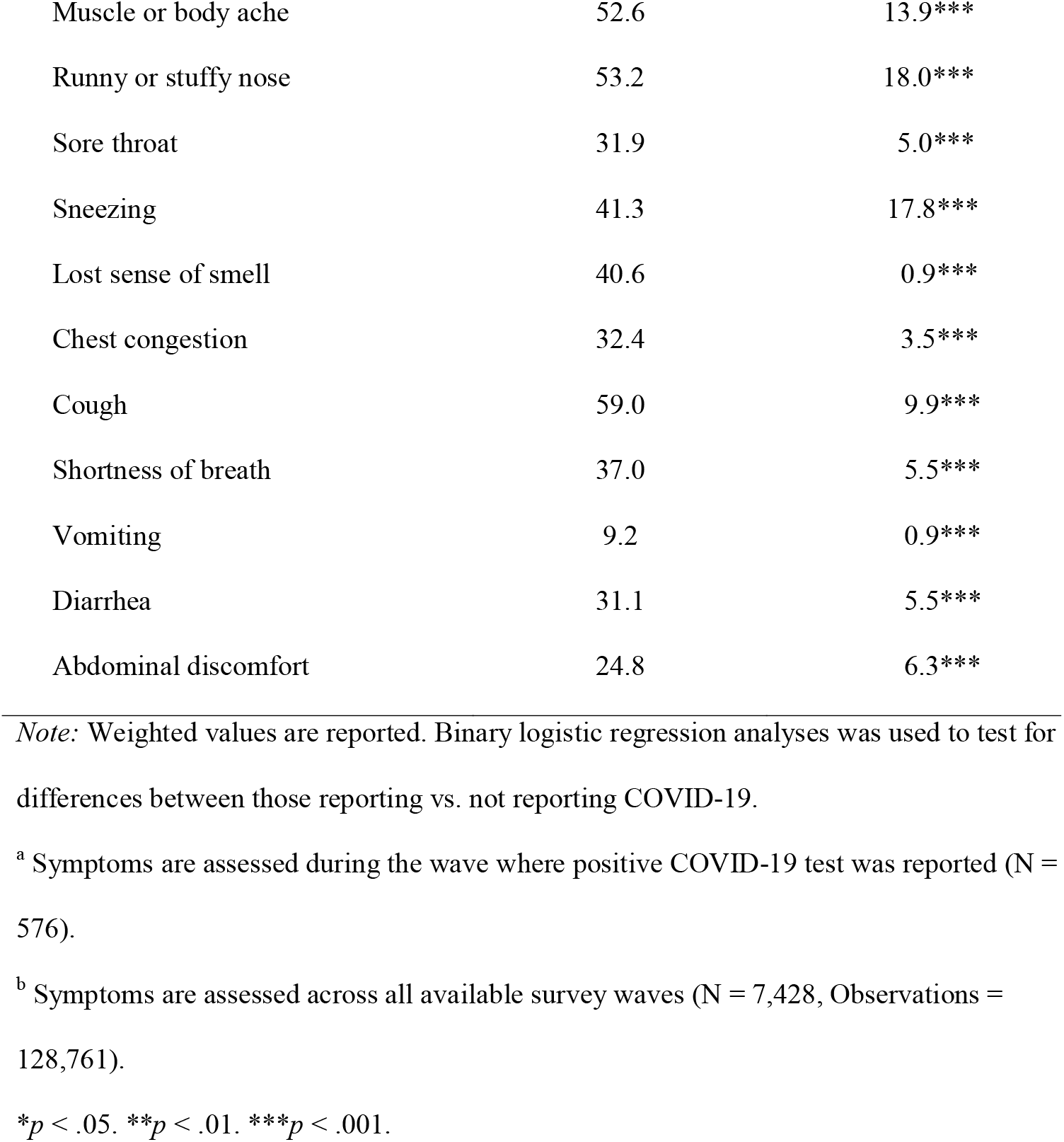
Demographic characteristics and symptoms experienced by participants reporting a positive COVID-19 test (N = 576) and remaining participants (N = 7,428) in the Understanding America Study.

### COVID-19 positive tests

Between April 1 2020 and February 15 2021 576 participants from the UAS sample reported that they had tested positive for COVID-19. On average 39.5% of symptoms were experienced in the past 7 days among those testing positive for COVID-19. The most common reported symptoms among those who tested positive were cough (59%), headache (58.9%), nose congestion (53.2%), muscle or body aches (52.6%), fever or high temperature (41.8%), and loss of smell (40.6%), as shown in Table 1. The prevalence of COVID-19 symptoms identified in the current study was comparable to that identified in high-quality studies where a positive COVID-19 test was confirmed via laboratory tests, as shown in Table S2.

The total estimated prevalence of COVID-19 across all survey waves of the UAS examined was 9.39% and the estimated prevalence of the disease in the US adult population over the same period was 9.42%. For further details of how prevalence estimates were derived see Section 1 of the Supplemental Materials. There was also a high degree of overlap in the time trend of COVID-19 cases in the UAS sample and the US adult population, as can be seen in Figure S1. The prevalence of COVID-19 across all UAS survey waves correlated strongly with the prevalence in the US population over the same two-week periods (r(21) = 0.93, p <.001), as shown in Figures S2 and S3.

### Fixed effects regression analysis

An initial examination of the descriptive trends in psychological distress tracked from before to after reporting a positive COVID-19 test indicted that distress rose and peaked in the wave when a positive COVID-19 test was initially reported and declined in the weeks that followed. This trend was observed in males and females, as illustrated in the unadjusted trends presented in Figure 1.

**Figure 1.**
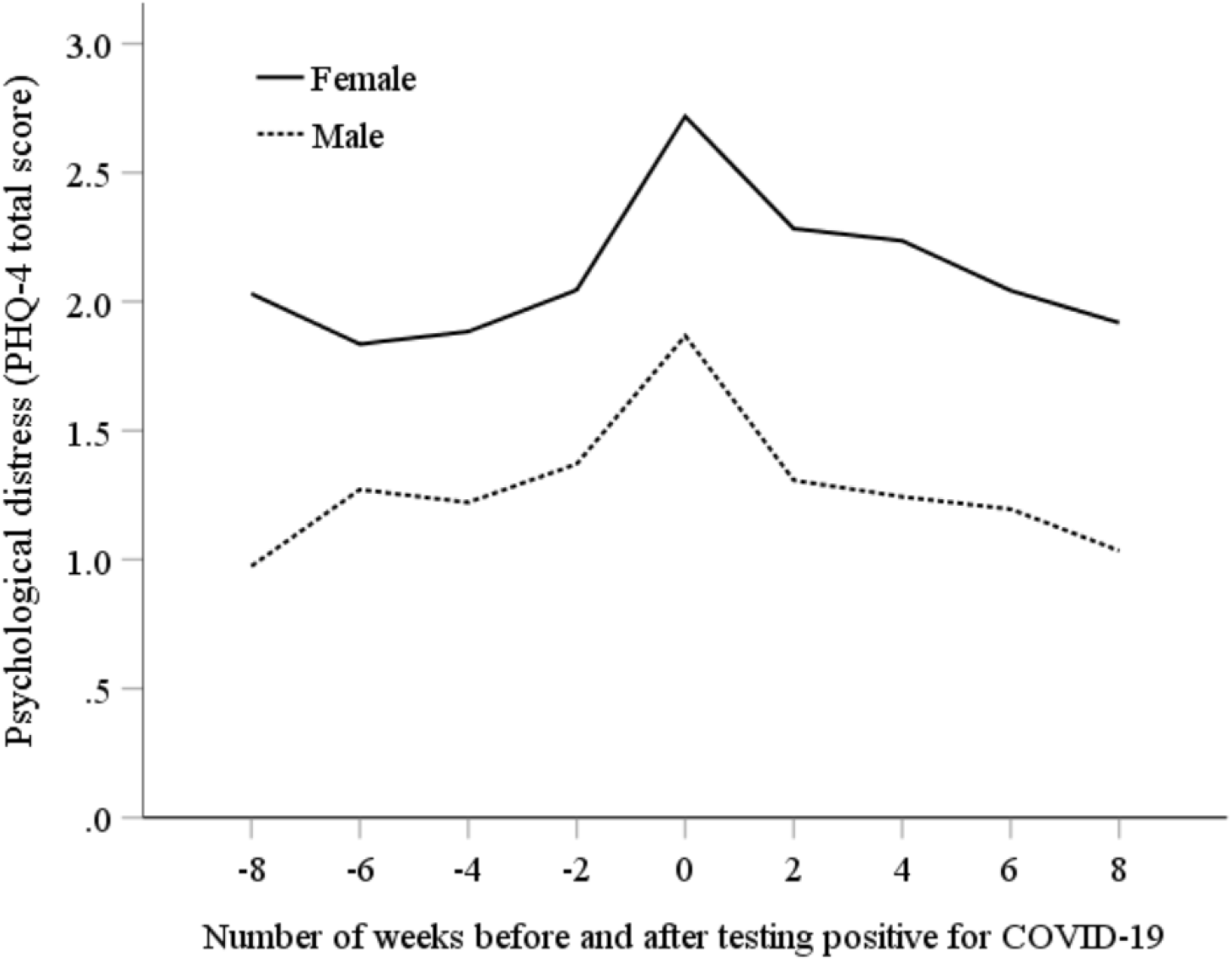
Trends in psychological distress as gauged by the PHQ-4 (range = 0-12) in the weeks before and after testing positive for COVID-19.

We found that testing positive for COVID-19 was associated with a 0.29 standard deviation (SD) increase in psychological distress in the same survey wave (B = 0.29, SE = 0.04, *p* <.001). This increase reflected a change from a predicted level of distress of -0.03 (95% CI: -0.03, -0.02) in non-COVID-19 waves to 0.26 (95% CI: 0.18, 0.35) within the survey wave where participants tested positive for COVID-19. In contrast, those who tested negative for COVID-19 experienced a very small 0.03 SD increase in distress (B = 0.03, SE = 0.01, *p* <.01). Lead effects of testing positive for coronavirus were non-significant indicating that distress did not increase substantially in advance of testing positive, as shown in Table 2. In contrast, there was evidence for a lag effect where testing positive for COVID-19 was associated with a 0.16 SD increase in distress in the next survey wave (B = 0.16, SE = 0.06, *p* <.01) two-weeks later. There was no evidence of significant associations between COVID-19 and distress beyond two weeks (lags of up to eight weeks / four survey waves tested). Further, the association between testing positive for COVID-19 and psychological distress was not moderated by sociodemographic factors or the presence of mental health conditions (see Table S3).

**Table 2.**
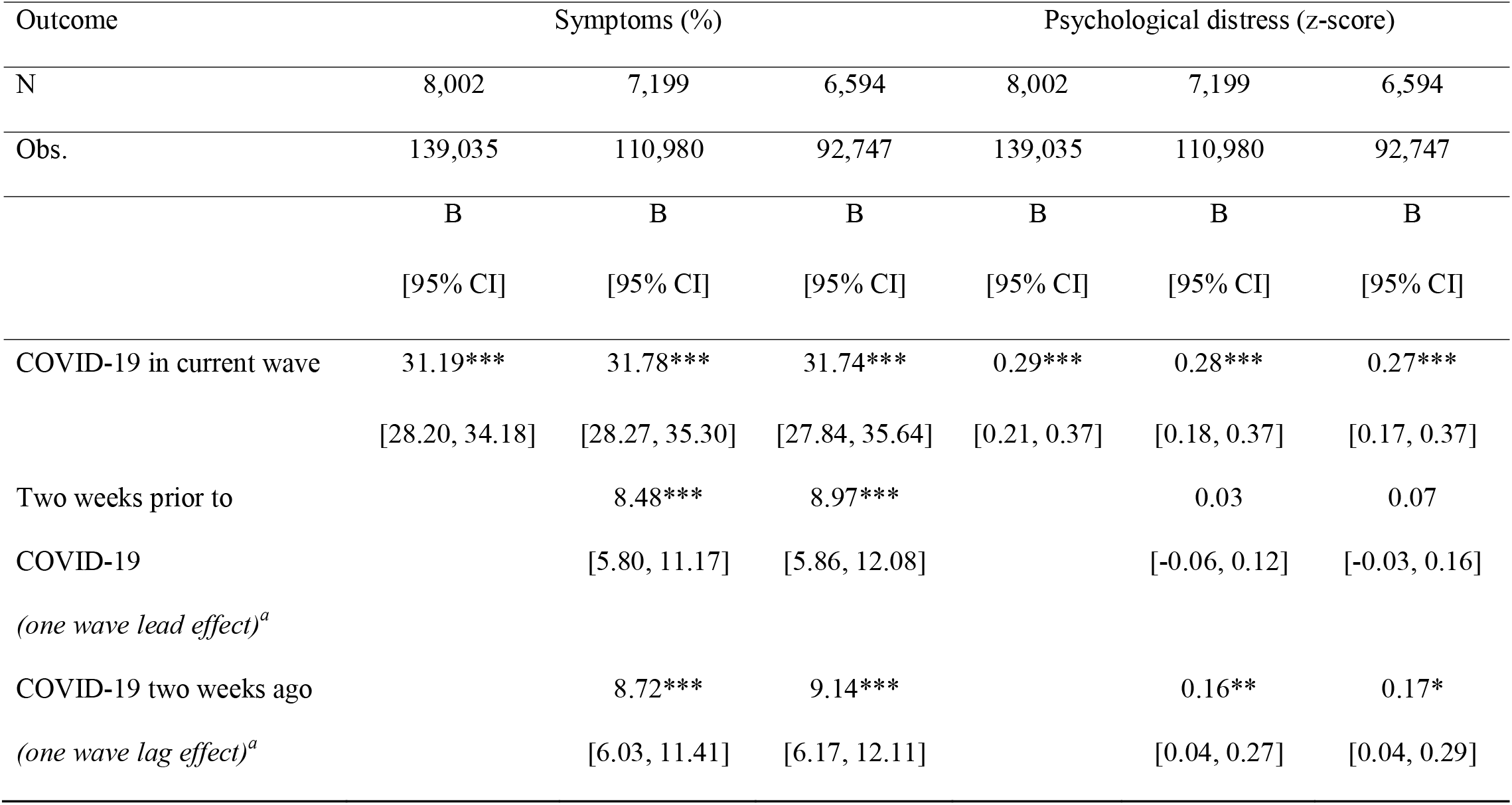
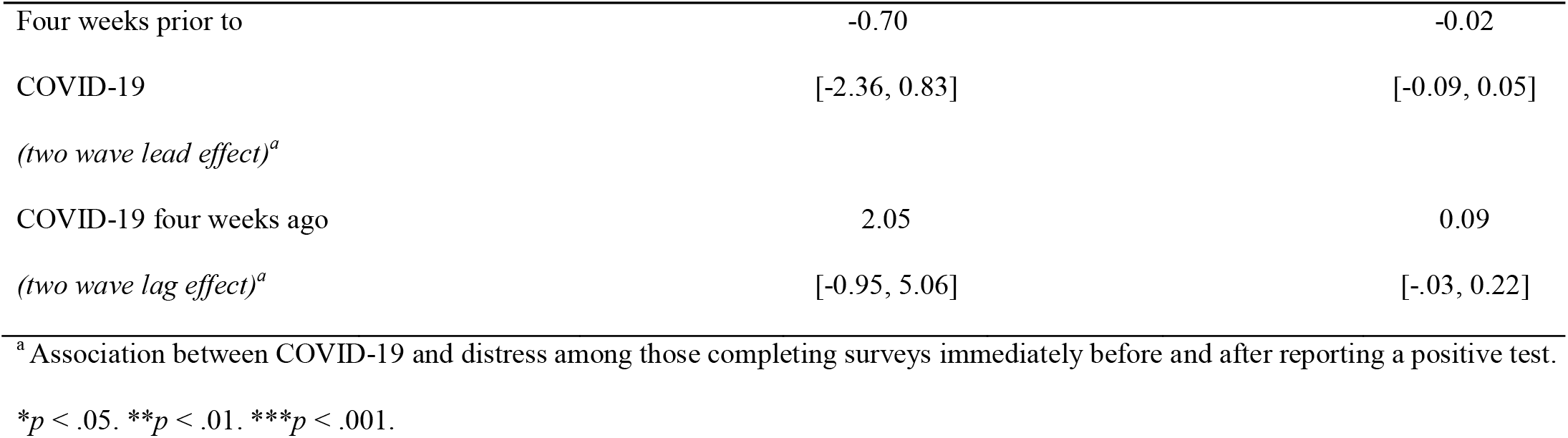
Fixed effects regression estimates of the association between testing positive for COVID-19 and changes in reported symptoms and psychological distress in the Understanding America Study.

Testing positive for COVID-19 was associated with a 31.2 percentage point (*p* < .001) increase in the percentage of COVID-19 symptoms from 8.1% (non-COVID-19 waves) to 39.3% (COVID-19 wave), as shown in Table 2. The percentage of symptoms reported increased by 8.5% (*p* <.001) from baseline levels in the two-week period prior to testing positive for COVID-19 and remained 8.7% (*p* <.001) above baseline levels in the two-week period after testing positive for the disease. There was no evidence of increases in symptom prevalence more than two weeks before or after the survey wave where the participant reported testing positive for COVID-19 (lags and leads of up to eight weeks / four survey waves tested).

### Mediation analysis

Changes in symptoms of COVID-19 were positively associated with changes in psychological distress in a fixed effects regression model. A change from experiencing no symptoms of COVID-19 to experiencing all symptoms assessed was associated with a 0.48 standard deviation increase in distress (B = 0.48, SE = 0.03, *p* <.001). In our mediation model, the indirect effect of COVID-19 symptoms (B = 0.15, SE = 0.01, p < .001) accounted for 52% of the association between testing positive for the disease and the increase in psychological distress (see Table 3). The direct effect of testing positive remained significant after adjustment for symptom levels (B = 0.14, SE = 0.04, p < .01). The indirect effect of symptoms (B = 0.08, SE = 0.02, p < .001) accounted for over half (52.8%) of the lagged effect of COVID-19 on distress two weeks later, and after accounting for symptom levels the relationship between infection and lagged increase in distress was non-significant. This finding indicates that once symptoms subsided, testing positive for COVID-19 was no longer significantly associated with increased distress.

**Table 3.** The role of COVID-19 symptoms as mediators of the association between testing positive for COVID-19 and psychological distress in the United States.

| Timing of positive<br>COVID-19 test | Psychological distress (z-score) |  |  |  |
| --- | --- | --- | --- | --- |
|  | Current wave |  | Previous wave |  |
|  | B | SE | B | SE |
| Total effect of COVID-19<sup>a</sup> | 0.29*** | 0.04 | 0.16** | 0.06 |
| Direct effect of COVID-19<sup>b</sup> | 0.14** | 0.04 | 0.07 | 0.06 |
| Indirect effect via<br>COVID-19 symptoms<sup>c</sup> | 0.15*** | 0.01 | 0.08*** | 0.02 |

*Note:* Psychological distress is standardized to have a mean of zero and standard deviation of one.
<sup>a</sup> Total increase in psychological distress associated with testing positive for COVID-19.
<sup>b</sup> Increase in psychological distress associated with testing positive for COVID-19 not explained by COVID-19 symptoms.
<sup>c</sup> Increase in psychological distress associated with testing positive for COVID-19 explained by COVID-19 symptoms.
\* $p < .05$ . \*\* $p < .01$ . \*\*\* $p < .001$

## Discussion

In this longitudinal population-based study, we used 23 biweekly waves of nationally representative longitudinal data to uncover evidence of changes in psychological distress in response to testing positive for COVID-19. Over the course of almost a full year of the pandemic 576 participants reported a positive COVID-19. We found that testing positive for COVID-19 was associated with an acute increase in distress levels (0.29 SD increase) after accounting for time-invariant unobserved heterogeneity using a fixed effects model. This spike in distress declined substantially within two weeks and was not significantly different to baseline levels after this point. The increase and then partial recovery in psychological distress after two weeks did not vary markedly across sociodemographic groups and was similar for those with/without a pre-existing mental health condition. Findings are consistent with proposals that those who contract COVID-19 may be at a greater risk of increased mental health symptoms compared to others living through the pandemic but not directly affected (Aknin et al., 2021). However, consistent with studies on mental health in the general population during the pandemic (Prati & Mancini, 2021; Robinson & Daly, 2020; Robinson et al., 2021), the present findings indicate that increases in distress attributable to infection are likely to be relatively transient and short-lived. Even after contracting a potentially deadly virus, the majority of the population appear to show resilience in mental health (van der Velden et al., 2021).

Because there have been concerns about the mental health consequences and long-term persistence of COVID-19 symptoms (e.g. Del Rio et al., 2020; Mahase, 2020) we examined changes in symptoms as a potential explanation for raised distress levels. In line with the overall results, symptoms increased during the wave of data collection when a positive COVID-19 test was first reported and declined rapidly thereafter. Further, those experiencing a greater number of COVID-19 symptoms showed the largest increases in distress and COVID-19 symptoms explained over half of the prospective association between testing positive for COVID-19 and experiencing raised distress.

The short-term increase in distress observed may be explained by the likely combination of physical symptoms (i.e. the direct effects of ill health on mental health) and initial worry over health having tested positive, the latter which presumably dissipates once symptoms have subsided (Taquet et al., 2021). Overall, our findings are consistent with cross-sectional studies linking COVID-19 symptoms to measures of mental health (Hyland et al., 2020; Wathelet et al., 2020). However, unlike previous research which relied on between-person estimates, the present research confirms that within-person changes in distress occur in response to a self-reported positive test for COVID-19. In the present study, once COVID-19 symptoms had dissipated at around two weeks from reporting a positive test, levels of distress returned to baseline levels. Although we found no evidence of long-term mental health consequences of COVID-19 in the present study, recent findings suggest that if a severe infection occurs (Taquet et al., 2021) or if physical symptoms are maintained over time (i.e. “long COVID”), there may be elevated levels of distress in this minority of the population (Del Rio et al., 2020; Sudre et al., 2021).

A key strength of the current study was the repeated longitudinal assessment which allowed the point at which a positive COVID-19 test was reported and associated changes in distress to be identified within a narrow time window in a large nationally representative sample of US adults. However, this study is limited in several respects. We relied on participant reports of a positive COVID-19 test rather than verified test results and self-reports are known to be vulnerable to bias. However, we conducted a number of analyses to estimate the likely accuracy of self-reports. First, the prevalence of COVID-19 symptoms among those reporting having tested positive for COVID-19 in the current study was comparable to that from large-scale studies that have used both self-report and objective verification of infection (Eythorsson et al., 2020; Wohl et al., 2021). The estimated cumulative prevalence of COVID-19 cases in the US population and the UAS sample were closely aligned (9.4% in each) and we observed strong concordance between the time trend of positive cases in the population and the UAS sample. In addition, we found that those who sought a COVID-19 test (presumably due to concerns about being infected) but tested negative showed minimal adverse change in their distress levels, which indicates that findings are unlikely to be explained by general concerns or worries about health as opposed to an actual COVID-19 infection. We also found no evidence that testing positive for COVID-19 predicted increased attrition or number of surveys completed in the present study. This suggests that bias in our assessment of confirmed cases may have been minimal.

Although we do not find evidence that testing positive for COVID-19 increases risk of dropout, it is likely that participants with the most severe infections were not included in the present study. Recent evidence suggest that an increased risk of common psychiatric disorders may be experienced chiefly by a subset of hospitalized patients experiencing encephalopathy (brain disorder, disease, or dysfunction) as a result of severe COVID-19 infection (Taquet et al., 2021; Taylor, Landry, Paluszek, Rachor, & Asmundson, 2020).

Therefore, the present findings provide information on the mental health consequences of testing positive for COVID-19 among the overall general population and not among those who developed COVID-19 and become critically ill or have suffered major adverse effects. It will therefore be important to continue to monitor long-term mental health outcomes on the population level and among those experiencing severe COVID-19 infections.

We examined a range of population demographics, including sub-groups who are at increased risk of becoming seriously ill after testing positive for COVID-19 (e.g. pre-existing physical health conditions, ethnic minorities) but found no evidence that changes in distress differed across demographics. Yet, while we utilized a large sample and a substantial portion of participants tested positive for the coronavirus, it remains possible that we were not well powered to detect small changes in long-term mental health among the population sub-groups examined. Furthermore, it may be the case that some population sub-groups experience worse mental health outcomes as a result of infection (e.g. those who become financially insecure due to being unable to work) and further examination of trends in mental health symptoms as a result of contracting COVID-19 is needed.

## Conclusions

Among a nationally representative sample of US adults, testing positive for COVID-19 is associated with an initial increase in psychological distress that diminishes quickly as symptoms subside. While COVID-19 may not produce lasting psychological distress among the majority of the general population it remains possible that a minority may suffer longer-term mental health consequences.

## Data Availability

The research data are distributed by the USC Dornsife Center for Economic and Social Research and available at https://uasdata.usc.edu/index.php

## Competing interests

ER has been a named investigator on research projects funded by the American Beverage Association and Unilever, but does not consider this funding a conflict of interest.

## Acknowledgements and Contribution

MD and ER designed research, MD analyzed data, & MR and ER performed research and wrote the paper.

## Acknowledgements

The project described in this paper relies on data from survey(s) administered by the Understanding America Study, which is maintained by the Center for Economic and Social Research (CESR) at the University of Southern California. The content of this paper is solely the responsibility of the authors and does not necessarily represent the official views of USC or UAS. The collection of the UAS COVID-19 tracking data is supported in part by the Bill & Melinda Gates Foundation and by grant U01AG054580 from the National Institute on Aging. However, these organizations bear no responsibility for the analysis or interpretation of the data.

## Ethical Approval

The UAS was approved by the University of Southern California human subjects committee internal review board (IRB) and informed consent was obtained from all participants.

## Notes

**Funding Statement**: This research received no specific grant from any funding agency, commercial or not-for-profit sector.

### Funding Statement

This research received no specific grant from any funding agency, commercial or not-for-profit sector.

### Author Declarations

Maynooth University Social Research Ethics Sub-Committee (SRESC). It is official policy of the SRESC that "ethical approval is not required for secondary use of anonymous data." which includes the two studies in this paper.

