## Supplemental Materials and Tables for "Acute and longer-term psychological distress associated with testing positive for COVID-19: longitudinal evidence from a population-based study of US adults"

* Corresponding author: Michael Daly

Address correspondence to:

Michael Daly Ph.D.

**Supplemental Materials Section 1:**

### Summary of method for estimating the prevalence of COVID-19 cases in the Understanding America Study (UAS) and the US population from April 1 2020 to February 15 2021

First, the prevalence of COVID-19 was estimated in each wave of the UAS. Survey weights were applied and the percentage of the sample reporting that they tested positive for COVID-19 since the last survey wave was examined. The sample and observations examined was identical to that reported in the main analyses (N = 8,002, Obs. = 139,035). Examining the percentage of those who took part in each survey who tested positive for COVID-19 produces a more accurate representation of the prevalence of positive cases in the UAS than simply taking the total number of cases (N = 576) and dividing this number by the total sample size (N = 8,002). This is because each wave of the UAS is weighted to represent the US population and therefore the percentage of positive cases in each individual survey wave is the figure of relevance when estimating prevalence in that wave and this also applies when producing cumulative estimates of prevalence by summing across waves.

Estimates of the percentage of the population testing positive for COVID-19 for each two-week period of the UAS survey can be seen in Figure S1. In total the cumulative prevalence of positive COVID-19 cases in the UAS was 9.39% between April 1 2020 and February 15 2021.

Next, to estimate the percentage of adults in the US population testing positive for COVID-19 we examined daily case numbers drawn from the Centers for Disease Control and Prevention COVID Data Tracker (CDC, 2021). Daily case numbers were calculated for each two-week period corresponding to the dates of the UAS surveys from April 1 2020 to February 15 2021. The number of cases reported for each two-week period was converted to a percentage of the US population by dividing case numbers for each period by the total US population, estimated as 330,086,374 at the end-point of the survey (https://www.census.gov/popclock/).

The total number of cases recorded by the CDC for each day of the pandemic includes those under 18 who have tended to be less likely to test positive for COVID-19 (Duca et al., 2020). Because a substantial portion of COVID-19 cases are missing information on demographics including age we estimated the impact of including those under 18 on our estimates using available data.

As of March 16 2021, the CDC had recorded 22,265,194 cases with age available. Based on this information the estimated percentage of the US population testing positive for COVID-19 was 6.75%. In contrast, using the same case figures the estimated prevalence of COVID-19 among those aged 18 and over was 7.66%. This meant that prevalence estimates that include those under 18 need to be inflated by a factor of 1.135 in order to more accurately represent the prevalence of positive tests in the adult population. As such, for each two-week period of the study the number of cases reported by the CDC in the population was inflated by this figure to estimate the percentage of those 18 and above who tested positive for COVID-19.

Using this methodology the estimated cumulative prevalence of positive COVID-19 tests among adults in the US population was 9.42% between April 1 2020 and February 15 2021. The correspondence between the UAS and US population estimates of the prevalence of positive COVID-19 cases is shown in Figures S1 to S3.

Table S1.

Number of participants and response rate among participants in the current study by survey wave from April 1 2020 to February 15 2021 (N = 8,002, Obs. = 139,035).

| Time-point | Survey period[a] | N | Response rate (%)[c] |
|---|---|---|---|
| Any | April 1 2020 – February 15 2021 | 8,002 | – |
| 1 | April 1 – 14 | 5,392 | 67.4 |
| 2 | April 15 – 28 | 6,198 | 77.4 |
| 3 | April 29 – May 12 | 6,301 | 78.7 |
| 4 | May 13 – 26 | 6,237 | 77.9 |
| 5 | May 27 – June 9 | 6,250 | 78.1 |
| 6 | June 10 – July 23 | 6,192 | 77.4 |
| 7 | June 24 – July 7 | 5,946 | 74.3 |
| 8 | July 8 – July 21 | 6,156 | 76.9 |
| 9 | July 22 – August 4 | 6,228 | 77.8 |
| 10 | August 5 – 18 | 6,081 | 76.0 |
| 11 | August 19 – September 1 | 6,103 | 76.3 |
| 12 | September 2 – 15 | 6,142 | 76.7 |
| 13 | September 16 – 29 | 5,962 | 74.5 |
| 14 | September 30 – October 13 | 5,931 | 74.1 |
| 15 | October 14 – 27 | 6,040 | 75.5 |
| 16 | October 28 – November 10 | 6,110 | 76.4 |

| | | | |
|---|---|---|---|
| 17 | November 11 – 24 | 5,926 | 74.0 |
| 18 | November 25 – December 8 | 5,888 | 73.6 |
| 19 | December 9 – 22 | 5,879 | 73.5 |
| 20 | December 23 – January 5 | 5,877 | 73.4 |
| 21 | January 6 – January 19 | 6,019 | 75.2 |
| 22 | January 20 – February 1 | 6,045 | 75.5 |
| 23 | February 2 – February 15 | 6,132 | 76.6 |

[a] Responses were permitted for an additional two weeks after the allotted two-week slot for each survey wave and a small portion of responses (max. ≈ 10%) of responses were made during this period and are included in the sample for each wave.

[b] Number of observations included in the current study for each survey wave.

[c] Indicates the percentage of the total number of participants included in the current study (N = 8,002) who have completed a survey in this wave.

Table S2.

Prevalence of COVID-19 symptoms reported among those who reported testing positive for COVID-19 in the Understanding America Study (UAS; N = 576) and those with confirmed positive COVID-19 tests in a population-based cohort from Iceland (N = 1,564) and a large-scale screening study in South Carolina (N = 1,116).

| | UAS[a] (N = 576) | Iceland[b] (N = 1,564) | South Carolina[c] (N = 1,116) |
|---|---|---|---|
| Symptom | % | % | % |
| Generalized symptoms | | | |
| Fever / raised temperature | 41.8 | 41.3 | 49.1 |
| Headache | 58.9 | 51.2 | 64.2 |
| Muscle or body ache | 52.6 | 54.6 | 53.7 |
| Upper respiratory | | | |
| Runny or stuffy nose | 53.2 | 33.3 | 45.4 |
| Sore throat | 31.9 | 33.7 | 48.8 |
| Sneezing | 41.3 | – | – |
| Lost sense of smell | 40.6 | 20.5 | 33.5 |
| Lower respiratory | | | |
| Chest congestion | 32.4 | – | – |
| Cough | 59.0 | 62.7[d] | 55.8 |
| Shortness of breath | 37.0 | 25.2 | 19.0 |
| Gastrointestinal | | | |

| | | | |
|---|---|---|---|
| Vomiting | 9.2 | 3.4 | 15.5 |
| Diarrhea | 31.1 | 13.7 | 16.3 |
| Abdominal discomfort | 24.8 | 11.1 | 11.7 |
| Average % of symptoms reported (all symptoms examined) | 39.5 | 31.8 | 37.5 |
| Average % of symptoms reported (symptoms common across all three studies) | 40.0 | 31.8 | 37.5 |

[a] Participants reported symptoms occurring in the past 7 days.

[b] Specific symptoms reported at symptom onset in a population-based cohort study assessed as part of a large-scale study aiming to characterize the occurrence of COVID-19 symptoms in the general population (Eythorsson et al., 2020).

[c] Symptoms reported during COVID-19 screening at the University of North Carolina Health Care (UNC Health) Respiratory Diagnostic Center (RDC) (Wohl et al., 2021).

[d] Combined reported prevalence of non-productive and productive cough at symptom onset.

Table S3.

Results from a series of fixed effects models examining interactions between demographic characteristics and testing positive for COVID-19 in predicting psychological distress in the Understanding America Study (N = 8,002, Obs. = 139,035).

| | Psychological distress (z-score) | | | |
|---|---|---|---|---|
| Timing of positive COVID-19 test | Current wave[a] | | Previous wave[b] | |
| Interaction effect (COVID-19*Variable) | β | SE | β | SE |
| 1. Ref. = 18-39 years | | | | |
| 1. Aged 40-59 | -0.08 | 0.09 | 0.07 | 0.13 |
| 1. Aged 60+ | 0.02 | 0.12 | 0.01 | 0.13 |
| 2. Female | 0.11 | 0.08 | 0.04 | 0.11 |
| 3. Ref. = White | | | | |
| 3. Hispanic | 0.20 | 0.11 | 0.19 | 0.15 |
| 3. Black | 0.15 | 0.17 | 0.36 | 0.21 |
| 3. Other race/ethnicity | 0.15 | 0.16 | 0.78 | 0.51 |
| 4. Ref. = Low income | | | | |
| 4. Medium income | -0.02 | 0.09 | -0.07 | 0.16 |
| 5. High income | 0.09 | 0.14 | -0.16 | 0.16 |
| 6. Physical health condition | -0.13 | 0.08 | 0.31 | 0.13 |
| 7. Mental health condition | 0.12 | 0.09 | 0.14 | 0.14 |

Note: p-value for statistical significance was set at < .01 due to multiple comparisons (20 interaction effects tested).

[a]Main effect of COVID-19 positive test and survey wave included in models but not shown.

[b] Main effect of COVID-19 positive test and lag and lead positive COVID-19 test effects and survey wave are included in all models but not shown.

** $p < .01$. *** $p < .001$.

Figure S1.

Estimated percentage of the US population (black line) and percentage of the Understanding America Study (UAS) sample testing positive for COVID-19 in each two-week survey period conducted from April 1 2020 to February 15 2021.

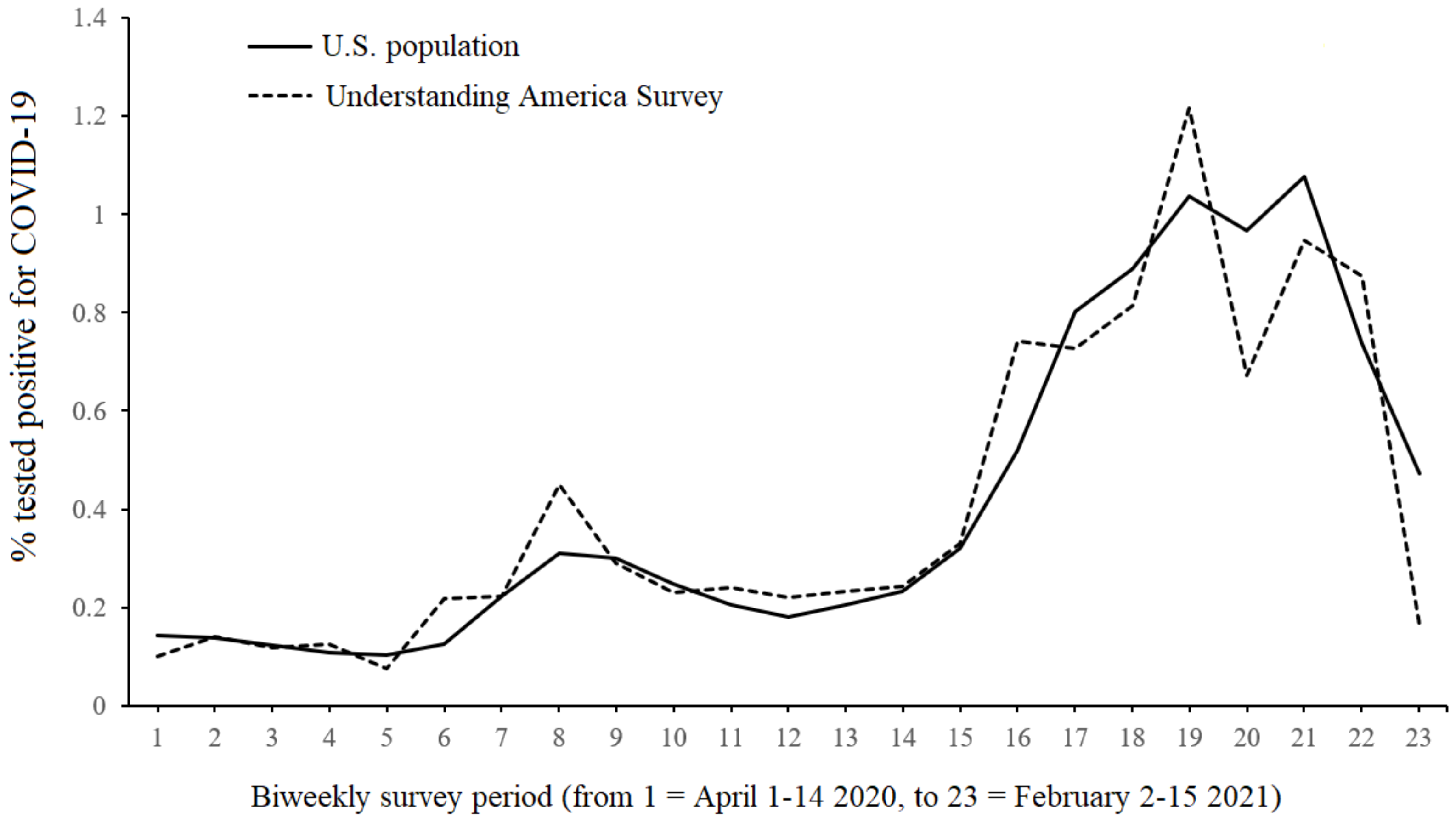

Figure S2.

Scatterplot of the estimated percentage of the US population and percentage of the Understanding America Study (UAS) sample testing positive for COVID-19 in each two-week survey period conducted from April 1 2020 to February 15 2021.

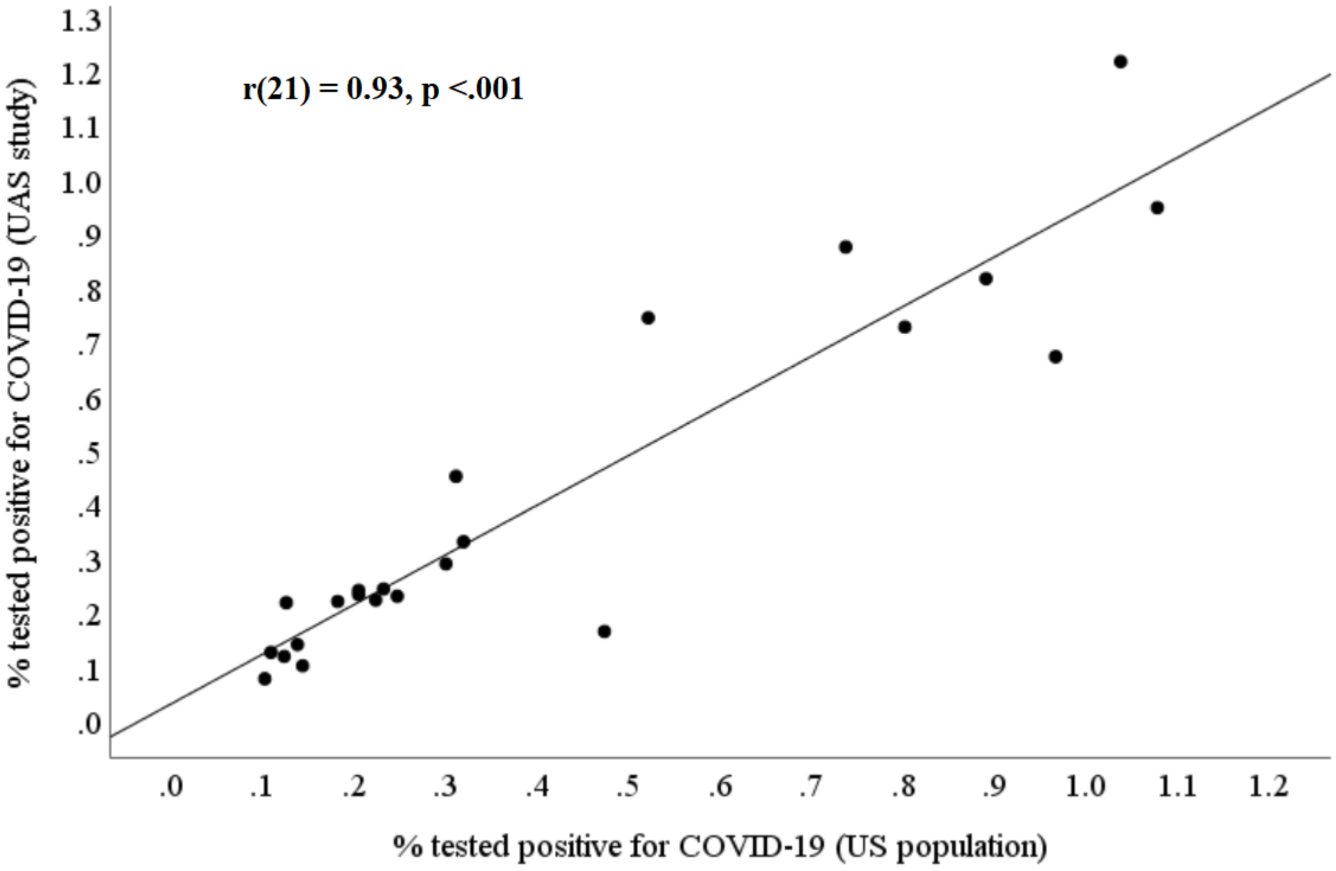

Figure S3.

Scatterplot of the natural logarithm of the percentage of the US population and the percentage of the Understanding America Study (UAS) sample testing positive for COVID-19 in each two-week survey period conducted from April 1 2020 to February 15 2021.

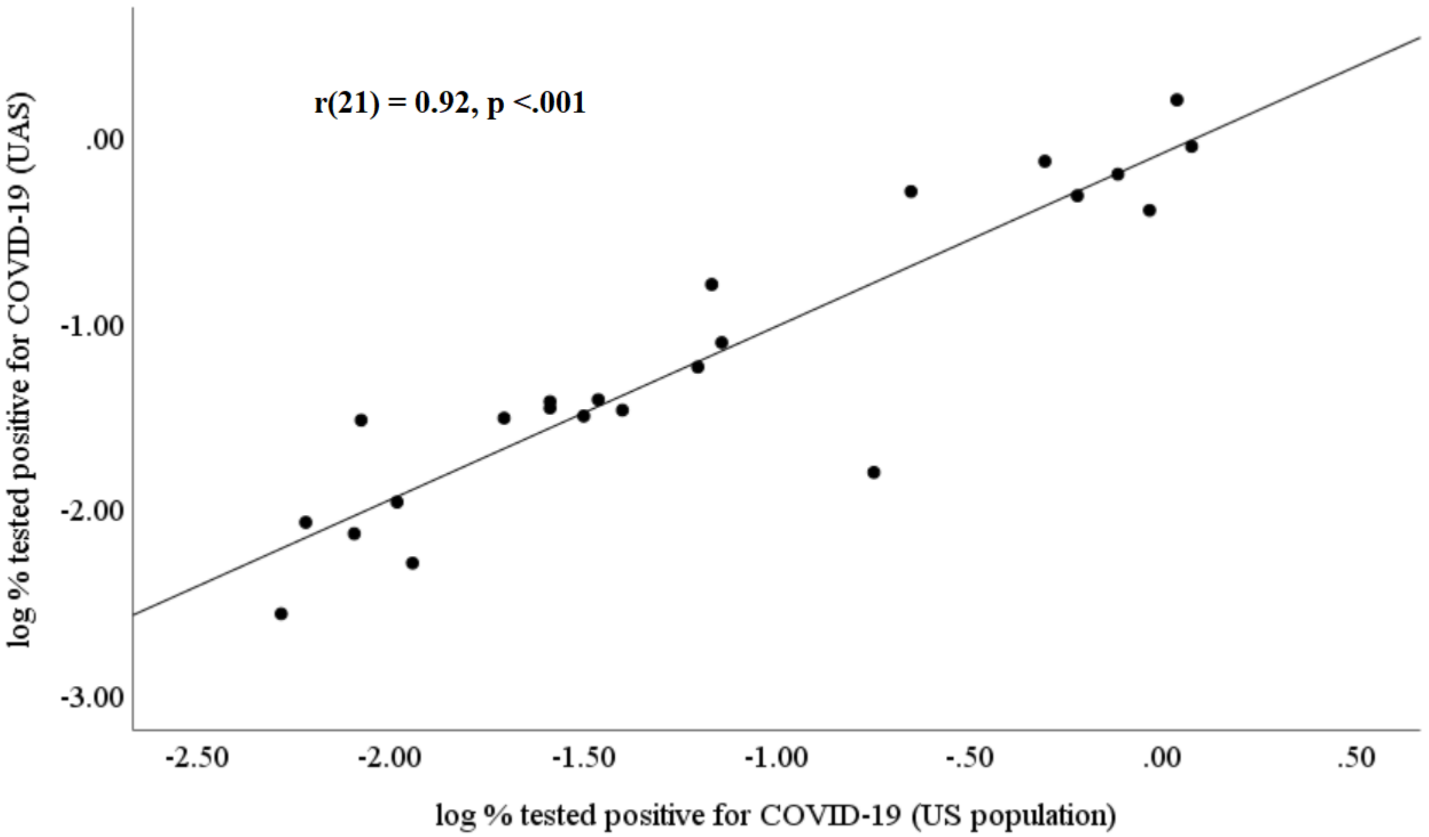
